# Limited user fees for adult HIV diagnosis and routine laboratory monitoring in the global IeDEA consortium: analysis in 42 countries

**DOI:** 10.64898/2025.12.05.25341715

**Authors:** Zhongzhe Pan, Ellen Brazier, Stephany N. Duda, Jeremy Ross, David J. Templeton, Sanjay Pujari, Lauren C. Zalla, Ronald M. Galiwango, Geoffrey Fatti, Gad Murenzi, Gabriela Carriquiry, Henri Chenal, Romanee Chaiwarith, Raynell Lang, Helen Byakwaga, Chido Chinogurei, Peter Ebasone, Carina Cesar, Oliver Ezechi, Aimee Freeman, Lameck O. Diero, Nicola van Dongen, Patricia Lelo, Sandra Wagner Cordoso, Ephrem Mensah, Kathleen McGinnis, Francesca Odhiambo, Ethel Rambiki, Christella Twizere, Marco Luque, Armel Poda, April D. Kimmel, the IeDEA consortium

## Abstract

**Introduction:** While HIV services have scaled up dramatically under universal treatment recommendations, availability of routine laboratory services for HIV diagnosis and management, and the extent to which user fees are charged for these services, are not well documented. We described site-level availability and user fees for adult HIV laboratory testing among 238 sites in 42 countries of the International epidemiology Databases to Evaluate AIDS (IeDEA) global consortium, and contextualized practices by site- and country-level characteristics, including country income level and receipt of Global Fund or PEPFAR support.

**Methods:** Availability of HIV-related laboratory services and charging of user fees, were examined via an IeDEA site assessment survey conducted September 2020–March 2021, with responses reflecting practices in 2019. Laboratory services examined included: HIV-1/HIV-2 antigen/antibody immunoassay test (HIV-1/2 Ag/Ab), HIV-1 p24 antigen test (p24), supplemental HIV-1/HIV-2 antibody differentiation immunoassay (HIV-1/2 Ab suppl), quantitative PCR for HIV viral load (VL), HIV-1 genotypic drug resistance testing (genotyping), and CD4 count testing (CD4). Availability was defined as provided on-site in the HIV clinic or within the same health facility. User fees were fees other than insurance co-pays paid for a given test, conditional on their on-site availability. We reported frequencies and percentages of site-level availability and user fees, overall and by site- and country-level characteristics.

**Results:** Most sites (88%) reported HIV-1/HIV-2 Ag/Ab availability on-site, while fewer than half of sites reported having p24 and HIV-1/2 Ab suppl available. Approximately half of sites reported VL and CD4 testing availability, while less than one-third of sites (28%) reported genotyping availability. The percentage of sites reporting user fees ranged from 6.5% (HIV-1/2 Ag/Ab) to 10.5% (CD4). A higher percentage of sites reporting user fees were from upper-middle income countries, countries receiving any Global Fund or any PEPFAR country support, and the Asia-Pacific region.

**Conclusions:** While most HIV care sites participating in IeDEA did not charge user fees for HIV-related laboratory tests in 2019, on-site availability of such tests was limited. Continued action to identify and address constraints in the routine provision of these laboratory services is critical for appropriate management of HIV.

## INTRODUCTION

HIV-related laboratory testing is important for diagnosis and monitoring treatment response for people with HIV [1], but can be costly [2]. A response has been user fees, defined as payments by patients for healthcare, medication or supplies at the point-of-service [3]. Typically imposed by facilities, user fees function as cost-recovery for services and, unlike insurance copays, are levied with all patients paying equal amounts [3, 4]. User fees can promote inequity for those with greater medical needs and increase disparities in access to healthcare services for those most vulnerable, such as those with HIV [5-9]. They can compromise care access, acting as a barrier to clinical appointments and routine laboratory monitoring, limiting antiretroviral therapy (ART) initiation and maintenance. Accordingly, user fees for HIV treatment and care were largely eliminated in the early 2000s [5], removing a key barrier to ART uptake and scale-up [5, 6, 10].

While the World Health Organization and other global health actors have discouraged the charging of user fees including for HIV treatment and care [5, 11-14], resource constraints have led to their re-introduction [15-18]. For example, even prior to the abrupt upheaval in the U.S. President’s Emergency Plan for AIDS Relief (PEPFAR) support and related HIV infrastructure in 2025 [19], sub-Saharan Africa faced unprecedented demands on its health systems given generally low levels of government health financing and gaps in HIV-related donor funding [20], which restricted availability and quality of HIV services [21, 22]. User fees were reintroduced for HIV diagnostic tests, routine laboratory monitoring, appointments and consultations, and treatment of opportunistic infections and comorbidities in parts of sub-Saharan Africa, such as in Nigeria in 2014 due to PEPFAR funding reductions [16]. They were also reported in the Democratic Republic of Congo and Guinea in 2019 [17], and over 25% of PEPFAR-supported countries reported user fees for HIV tests and services in 2020 [18].

Little is known about the landscape of user fees for HIV-related care globally, particularly in the universal treatment era [23]. We describe the availability and charging of user fees for adult HIV-related diagnosis and laboratory testing among HIV treatment and care sites participating in the International epidemiology Databases to Evaluate AIDS (IeDEA) global research consortium, and contextualize these practices by considering site- and country-level characteristics.

## METHODS

This study was designated a non-human subjects operational/quality improvement project by the Vanderbilt University Medical Center Institutional Review Board (#200013). Informed consent was not required because the study collected site-level data only and human subjects were not involved.

### Data

Data on laboratory testing services and user fees came from a survey completed by HIV treatment and care clinics participating in IeDEA in 2020, including sites in 42 countries across seven geographic regions: the Asia-Pacific; the Caribbean, Central America, and South America; North America; Central Africa; East Africa; Southern Africa; and West Africa. As described elsewhere [24], the survey explored site-level characteristics and HIV-related service availability during 2019, with survey responses reflecting routine clinic services and practices prior to the SARS-CoV-2 pandemic. The survey was conducted between September 2020 and March 2021 via online REDCap surveys [25, 26] or paper questionnaires distributed to 238 facilities providing adult, adolescent, and/or pediatric HIV treatment and care. Completed by clinic staff involved in HIV care, the survey had a response rate of 95% [24]. The survey examined the clinic population served (urban vs. rural; children, adolescents or adults); the availability of various HIV diagnosis and laboratory monitoring services—within the HIV clinic, elsewhere within the health facility, off-site via referral, not available—and the charging of user fees for these services.

Data also included publicly available country-level characteristics for 2019. World Bank country-level income level was defined as low, low-middle, middle-high, or high [27]. Country-level PEPFAR support was defined as any country with a country operational plan [28] and Global Fund to Fight AIDS, Tuberculosis and Malaria (Global Fund) support was any country with a Global Fund allocation [29]. Data on national HIV prevalence (<1%, >1–5%, >5%) came from UNAIDS [30].

### Analytic sample

The analytic sample included HIV treatment and care clinics responding to the IeDEA survey and serving adults. We excluded sites providing services only to children given clinical practice differences for pediatric patients.

### Survey measures

The survey examined on-site availability (i.e., provided in the HIV clinic, within the same health facility) of tests for HIV diagnosis and routine HIV monitoring. HIV diagnostic tests included HIV-1 p24 antigen test for acute HIV-1 infection (p24), HIV-1/HIV-2 antigen/antibody immunoassay test for established HIV infection (HIV-1/2 Ag/Ab), and supplemental HIV-1/HIV-2 antibody differentiation immunoassay (HIV-1/2 Ab suppl). Laboratory tests for routine monitoring included quantitative PCR for HIV viral load and HIV-1 genotypic drug resistance testing (genotyping). The survey also examined the availability of CD4 cell count tests [31].

User fees were defined in the survey as fees other than insurance co-pays paid by service users and for a given laboratory test. A given test was classified as having user fees, not having user fees, or unknown (do not know or missing response).

### Statistical Methods

We report frequencies and percentages of site-level availability of HIV-related laboratory tests. We also report frequencies and percentages of user fees for each test, conditional on their on-site availability, overall and by IeDEA region, clinic population urbanicity, and country-level characteristics (income group, any Global Fund support, any PEPFAR country support, HIV prevalence). Analyses were performed using SAS 9.4 (SAS Institute, Cary, NC).

## RESULTS

### Site characteristics

Of 238 IeDEA sites surveyed, 199 (84%) provided adult HIV care and were included in the analysis. The majority of sites were located in East Africa (37%), followed by the Asia-Pacific (19%), North America (15%), Southern Africa (13%) and Central Africa (10%), with fewer sites in the Caribbean, Central and South America region (4%) and West Africa (3%). Most sites served urban populations (64%), were in countries receiving any Global Fund allocations (72%) and any PEPFAR country support (69%), and were in lower-middle income countries (37%), followed by high income (26%), low income (25%), and upper-middle income countries (12%).

### Availability of services

The reported availability of laboratory testing varied widely (**Table 1**). The majority of sites (92%) reported HIV-1/HIV-2 Ag/Ab availability, but fewer than two in five sites reported p24 (37%) and HIV-1/2 Ab suppl (40%) availability. Viral load availability was reported by approximately half of sites (50%), as was CD4 cell count testing (53%). Genotyping was available at 30% of sites. Apart from HIV-1/HIV-2 Ag/Ab, which had near-universal availability, sites reporting availability of laboratory testing were predominantly from the Asia-Pacific or North American regions, served urban populations, and were in high- or upper-middle income countries, countries not receiving any Global Fund or any PEPFAR country support, and countries having HIV prevalence <1%.

**Table 1.** Summary of availability and user fees for HIV-related laboratory tests in the global IeDEA consortium.

| Characteristic | All Sites, N (%)<sup>1</sup> | Diagnostic Tests |  |  |  |  |  | Routine Management Tests |  |  |  |  |  |
| --- | --- | --- | --- | --- | --- | --- | --- | --- | --- | --- | --- | --- | --- |
|  |  | p24, N (%) |  | HIV-1/2 Ag/Ab, N (%) |  | HIV-1/2 Ab suppl, N (%) |  | Viral load, N (%) |  | Genotyping, N (%) |  | CD4, N (%) |  |
|  |  | Avail<sup>2</sup> | UF<sup>3</sup> | Avail<sup>2</sup> | UF<sup>3</sup> | Avail<sup>2</sup> | UF<sup>3</sup> | Avail<sup>2</sup> | UF<sup>3</sup> | Avail<sup>2</sup> | UF<sup>3</sup> | Avail<sup>2</sup> | UF<sup>3</sup> |
| Overall | 199 | 73 (37) | 6 (8) | 184 (92) | 12 (7) | 80 (40) | 8 (10) | 100 (50) | 8 (8) | 59 (30) | 4 (7) | 105 (53) | 11 (11) |
| IeDEA Region |  |  |  |  |  |  |  |  |  |  |  |  |  |
| Asia-Pacific | 37 (19) | 27 (73) | 4 (15) | 37 (100) | 6 (16) | 27 (73) | 5 (19) | 33 (89) | 5 (15) | 25 (68) | 2 (8) | 27 (73) | 6 (22) |
| Central Africa | 20 (10) | 4 (20) | 0 (0) | 17 (85) | 3 (17) | 6 (30) | 1 (17) | 7 (35) | 2 (29) | 0 (0) | - | 3 (15) | 3 (100) |
| CCASAnet | 7 (4) | 3 (43) | 0 (0) | 7 (100) | 0 (0) | 1 (14) | 0 (0) | 4 (57) | 0 (0) | 3 (43) | 0 (0) | 5 (71) | 0 (0) |
| East Africa | 74 (37) | 11 (15) | 0 (0) | 65 (88) | 1 (2) | 8 (11) | 0 (0) | 9 (12) | 0 (0) | 2 (3) | 0 (0) | 31 (42) | 1 (3) |
| NA-ACCORD | 30 (15) | 23 (77) | 1 (4) | 28 (93) | 1 (4) | 28 (93) | 1 (4) | 29 (97) | 1 (3) | 24 (80) | 1 (4) | 24 (80) | 1 (4) |
| Southern Africa | 25 (13) | 4 (16) | 1 (25) | 24 (96) | 1 (4) | 4 (16) | 1 (25) | 13 (52) | 0 (0) | 4 (16) | 0 (0) | 9 (36) | 0 (0) |
| West Africa | 6 (3) | 1 (17) | 0 (0) | 6 (100) | 0 (0) | 6 (100) | 0 (0) | 5 (83) | 0 (0) | 1 (17) | 1 (100) | 6 (100) | 0 (0) |
| Site Population Urbanicity |  |  |  |  |  |  |  |  |  |  |  |  |  |
| Urban | 127 (64) | 64 (50) | 10 (8) | 122 (96) | 5 (8) | 72 (57) | 7 (10) | 90 (71) | 8 (9) | 56 (44) | 4 (7) | 83 (65) | 10 (12) |
| Rural | 68 (34) | 9 (13) | 2 (3) | 58 (85) | 1 (11) | 7 (10) | 1 (14) | 9 (13) | 0 (0) | 3 (4) | 0 (0) | 19 (28) | 1 (5) |
| Country Income Group |  |  |  |  |  |  |  |  |  |  |  |  |  |
| Low | 49 (25) | 5 (10) | 0 (0) | 41 (84) | 2 (5) | 6 (12) | 0 (0) | 9 (18) | 0 (0) | 0 (0) | - | 17 (35) | 1 (6) |
| Lower-Middle | 74 (37) | 15 (20) | 2 (13) | 69 (93) | 5 (7) | 21 (28) | 4 (19) | 23 (31) | 5 (22) | 5 (7) | 2 (40) | 37 (50) | 6 (16) |
| Upper-Middle | 24 (12) | 12 (50) | 3 (25) | 7 (100) | 4 (17) | 9 (38) | 3 (33) | 20 (83) | 2 (10) | 13 (54) | 1 (8) | 15 (63) | 3 (20) |
| High | 52 (26) | 41 (79) | 1 (2) | 50 (96) | 1 (2) | 44 (85) | 1 (2) | 48 (92) | 1 (2) | 41 (79) | 1 (2) | 36 (69) | 1 (3) |
| Country Global Fund Support |  |  |  |  |  |  |  |  |  |  |  |  |  |
| Supported | 143 (72) | 28 (20) | 11 (9) | 130 (90) | 11 (9) | 34 (24) | 7 (21) | 48 (34) | 7 (15) | 14 (10) | 3 (21) | 65 (46) | 10 (15) |
| Not Supported | 56 (28) | 45 (80) | 1 (2) | 54 (96) | 1 (2) | 46 (82) | 1 (2) | 52 (93) | 1 (2) | 45 (80) | 1 (2) | 40 (71) | 1 (3) |
| Country PEPFAR Support |  |  |  |  |  |  |  |  |  |  |  |  |  |
| Supported | 137 (69) | 27 (20) | 4 (15) | 124 (90) | 10 (8) | 30 (22) | 6 (20) | 43 (31) | 6 (14) | 12 (9) | 3 (25) | 61 (45) | 9 (15) |
| Not Supported | 62 (31) | 46 (74) | 2 (4) | 60 (97) | 2 (3) | 50 (80) | 2 (4) | 57 (92) | 2 (4) | 47 (76) | 1 (2) | 44 (71) | 2 (5) |
| Country HIV Prevalence |  |  |  |  |  |  |  |  |  |  |  |  |  |
| <1% | 67 (34) | 47 (70) | 4 (9) | 65 (97) | 5 (7) | 52 (78) | 4 (8) | 59 (88) | 4 (7) | 45 (67) | 2 (4) | 52 (78) | 5 (10) |
| 1-5% | 66 (33) | 13 (20) | 0 (0) | 59 (89) | 3 (5) | 15 (23) | 1 (7) | 16 (24) | 2 (13) | 3 (5) | 1 (33) | 21 (32) | 3 (14) |
| 5-100% | 57 (29) | 6 (11) | 1 (17) | 51 (90) | 2 (4) | 6 (11) | 1 (17) | 16 (28) | 0 (0) | 4 (7) | 0 (0) | 25 (44) | 1 (4) |

Abbreviations: *IeDEA* = International epidemiology Databases to Evaluate AIDS, *Avail* = available, *UF* = user fees, *p24* = HIV-1 p24 antigen test for acute HIV-1 infection, *HIV-1/2 Ag/Ab* = HIV-1/HIV-2 antigen/antibody immunoassay test for established HIV infection, *HIV-1/2 Ab suppl* = supplemental HIV-1/HIV-2 antibody differentiation immunoassay, *viral load* = quantitative PCR for HIV viral load, *genotyping* = HIV-1 genotypic drug resistance testing, *CD4* = CD4 cell count test, *CCASAnet* = Caribbean, Central and South America network for HIV Epidemiology, *NA-ACCORD* = North American AIDS Cohort Collaboration on Research and Design, *PEPFAR* = U.S. President's Emergency Plan for AIDS Relief.
<sup>1</sup> The percentages reported are column percentages of all sites. Column percentages may not sum to one because we do not report sites with missing information.
<sup>2</sup> Availability is defined as services accessible in the HIV clinic or within the same health facility. Percentages reported are the number of sites with services available divided by all sites.
<sup>3</sup> User fees are defined as services reported as charging a user fee. Percentages reported are the number of sites reporting user fees for a given service divided by the number of sites with the service available.

### User fees

Few sites (7%) reported user fees for at least one HIV laboratory test. The percentage reporting user fees ranged from 7% (HIV-1/2 Ag/Ab and genotyping) to 11% (CD4) (Table 1). Except for genotyping, a higher percentage of sites reporting user fees were from the Asia-Pacific region (15–22% of sites depending on laboratory test type), Central Africa (17–100%) or Southern Africa (25%). Across all laboratory tests, as many as 40% of sites in lower-middle countries and 33% in upper-middle income countries reported user fees, while as many as 25% of sites in countries receiving any Global Fund support or any PEPFAR country support reported fees.

## DISCUSSION

Addressing a gap in data about user fee charges for HIV-related tests and services globally, the current study found that over 90% of HIV care sites in a diverse global cohort did not report user fees for HIV laboratory tests. However, there was considerable variation by country income status and sources of financing for HIV treatment and care.

We found limited availability of critical HIV laboratory tests. Fewer than half of sites reported on-site availability for HIV diagnosis (p24, HIV-1/2 Ab suppl) and treatment management (viral load, genotyping, CD4), suggesting test availability and accessibility may be a barrier to diagnosis, treatment and care. Laboratory test availability via integration of HIV-related services, such as point-of-care viral load testing, is effective in improving HIV management [32]. However, establishing such service delivery models can be costly: a budget impact analysis in South Africa found national scale-up of point-of-care viral load testing could require an additional $50 million annually [33]. While user fees to offset the on-site service costs could positively affect the availability of healthcare services [34], it could also generate financial barriers for accessing services [7].

A higher percentage of sites in upper-middle income countries reported user fees, particularly for HIV diagnostic testing. A possible explanation—given ongoing and historical funding gaps and donor funding that has not kept up with inflation [20]—is that these countries have faced challenges transitioning from reliance on external donors to sustaining the HIV response with domestic funds [35]. User fees could compensate for funding shortages. Notably, a higher percentage of sites from countries receiving any Global Fund or PEPFAR country support reported user fees. A possible explanation is that the external support may not fully offset site-level funding gaps resulting in a need to charge user fees.

Differences emerged across IeDEA geographic regions. Higher percentages of sites in the Asia-Pacific region reported user fees for almost all laboratory testing services examined. This region has the second largest population with HIV and approximately one-quarter of new infections globally [36]. Yet HIV resources are insufficient, with an estimated 60% funding gap in 2024 for meeting global targets to end the HIV epidemic[37]. Given approximately 85% of total financial resources for HIV diagnosis, treatment, and care there come from domestic funders [37], it is possible that these sites may implement user fees to offset costs. In contrast, most sites in IeDEA’s North America region are in the US, where the federal Ryan White HIV/AIDS Program and coverage through the Affordable Care Act and Medicaid expansion may reduce or eliminate fees for eligible patients [38]. However, current threats to the ongoing funding of such programs in the US, may result in decreasing coverage and the implementation of user fees to cover program costs.

Our study is among the first to describe the landscape of user fees across a globally diverse cohort of HIV clinics in countries with different health systems, income levels, and sources of HIV funding. While few sites reported user fees, shortages of HIV financing may play a role in user fees. Indeed, a study conducted in Nigeria found that 96% of sites re-introduced user fees to in response to reductions in PEPFAR support in 2014 [16]. Our study also provides context to literature finding an association between the introduction of user fees and HIV-related healthcare utilization and outcomes, such as reduced HIV diagnosis [10, 39], worsened new patient enrolment and decreased ART prescriptions [7]. Collectively, this suggests that the abrupt withdrawal of PEPFAR funding in 2025 may trigger the re-introduction of user fees globally and, thus, worsen HIV-related healthcare utilization [7]; ongoing surveillance of user fees and their impact on outcomes is urgently needed.

### Limitations

First, while survey questions were intended to capture laboratory services availability and user fees in 2019, prior to the COVID-19 pandemic period, responses could have been subject to recall bias. Second, we relied on site-level self-report, which may be subject to social desirability bias (i.e., respondents may not wish to report user fees) and other information biases (e.g., respondents may not have fully identified all types of user fees occurring for patients). Third, most participating sites served urban populations, and urban sites may be better resourced than peripheral rural clinics. Accordingly, our findings may overestimate test availability. Finally, we did not collect information on the amount of user fees charged, and cannot assess associated financial barriers for HIV diagnosis, treatment and care.

## Conclusions

We found limited availability of critical HIV-related laboratory tests. While most HIV care sites globally did not report user fees in 2019, our findings suggest that continued funding of comprehensive HIV treatment and care services is critical to preserving progress toward ending the HIV epidemic.

## Competing Interests

The authors declare no competing interests.

## Author Contributions

A.D.K. conceptualized the research. Z.P. and A.D.K. designed the analysis. Z.P. conducted the analysis. Z.P., E.B., and A.D.K. drafted the manuscript. Z.P., E.B., S.D., J.R., D.T., S.P., L.C.Z., R.M.G, G.F., G.M., G.C., H.C., R.C., R.L., H.B., C.C., P.E., C.C., O.E., A.F., L.O.D., N.V.D., P.L., S.W.C., E.M., K.M., F.O., E.R., C.T., M.L., A.P., and A.D.K. interpretated the data and critically reviewed and revised the manuscript. All authors approved the final manuscript.

## Data Availability

The data that support the findings of this study are available from the corresponding author upon reasonable request. A request to access the survey data from the IeDEA consortium for research purposes may be made through submission of a formal concept proposal, which is available at https://www.iedea.org/.

## Funding

The International epidemiology Databases to Evaluate AIDS (IeDEA) is supported by the U.S. National Institutes of Health’s (NIH) National Institute of Allergy and Infectious Diseases, the *Eunice Kennedy Shriver* National Institute of Child Health and Human Development, the National Cancer Institute, the National Institute of Mental Health, the National Institute on Drug Abuse, the National Heart, Lung, and Blood Institute, the National Institute on Alcohol Abuse and Alcoholism, the National Institute of Diabetes and Digestive and Kidney Diseases, and the Fogarty International Center: ***Asia-Pacific***, U01AI069907; ***CCASAnet***, U01AI069923; ***Central Africa***, U01AI096299; ***East Africa***, U01AI069911; ***NA-ACCORD***, U01AI069918; ***Southern Africa***, U01AI069924; ***West Africa***, U01AI069919. Informatics resources are supported by the Harmonist project, R24AI124872. This work is solely the responsibility of the authors and does not necessarily represent the official views of any of the institutions mentioned above.

This publication is the result of funding in whole or in part by the NIH. It is subject to the NIH Public Access Policy. Through acceptance of this federal funding, NIH has been given a right to make this manuscript publicly available in PubMed Central upon the Official Date of Publication, as defined by NIH.

## Notes

### Competing Interest Statement

The authors have declared no competing interest.

